# Optimising biomarkers for accurate ependymoma diagnosis, prognostication and stratification within International Clinical Trials: A BIOMECA study

**DOI:** 10.1101/2022.11.24.22282659

**Authors:** Rebecca J. Chapman, David R. Ghasemi, Felipe Andreiuolo, Valentina Zschernack, Arnault Tauziede Espariat, Francesca Buttarelli, Felice Giangaspero, Jacques Grill, Christine Haberler, Simon M.L. Paine, Ian Scott, Thomas S. Jacques, Martin Sill, Stefan Pfister, John-Paul Kilday, Pierre Leblond, Maura Massimino, Hendrik Witt, Piergiorgio Modena, Pascale Varlet, Torsten Pietsch, Richard G. Grundy, Kristian W. Pajtler, Timothy A. Ritzmann

**Author notes:** **Corresponding author** Timothy Ritzmann, Children’s Brain Tumour Research Centre, Biodiscovery Institute, University of Nottingham, Nottingham, UK. On Behalf of the Biomarkers of Ependymoma in Childhood and Adolescence (BIOMECA) Consortium. These authors contributed equally to senior authorship (Kristian W. Pajtler, Timothy A. Ritzmann). **AUTHORSHIP** Experimental design: RJC, FA, FG, JG, CH, JPK, PB, MM, HW, PM, TP, PV, RGG, KWP, TAR Implementation: RJC, FA, FB, ATE, CH, PM, TP, PV, HW, SMLP, IS, TJ, TAR Data analysis: RJC, DRG, TAR, VZ, FA, SMLP, ATE, CH, PM, PV, MS Data interpretation: RJC, DRG, TAR, RGG, TP, KWP Manuscript preparation: RJC, TAR, DRG, RGG, TP, KWP.

## Abstract

**Background:** Accurate identification of brain tumour molecular subgroups is increasingly important. We aimed to establish the most accurate and reproducible ependymoma subgroup biomarker detection techniques, across 147 cases from International Society of Pediatric Oncology (SIOP) Ependymoma II trial participants, enrolled in the pan-European “Biomarkers of Ependymoma in Children and Adolescents (BIOMECA)” study.

**Methods:** Across six European BIOMECA laboratories we evaluated epigenetic profiling (DNA methylation array); immunohistochemistry (IHC) for nuclear p65-RELA, H3K27me3, and Tenascin-C; copy number analysis via FISH and MLPA (1q, *CDKN2A*), and MIP and DNA methylation array (genome-wide copy number evaluation); analysis of *ZFTA-* and *YAP1*-fusions by RT-PCR and sequencing, Nanostring and break-apart FISH.

**Results:** DNA Methylation profiling classified 65.3% (n=96/147) of cases as EPN-PFA and 15% (n=22/147) as ST-ZFTA fusion-positive. Immunohistochemical loss of H3K27me3 was a reproducible and accurate surrogate marker for EPN-PFA (sensitivity 99-100% across three centres). IHC for p65-RELA, FISH, and RNA-based analyses effectively identified *ZFTA-* and *YAP1-* fused supratentorial ependymomas. Detection of 1q gain using FISH exhibited only 57% inter-centre concordance and low sensitivity and specificity whilst MIP, MLPA and DNA methylation-based approaches demonstrated greater accuracy.

**Conclusions:** We confirm, in a prospective trial cohort, that H3K27me3 immunohistochemistry is a robust EPN-PFA biomarker. Tenascin-C should be abandoned as a PFA marker. DNA methylation and MIP arrays are effective tools for copy number analysis of 1q gain, 6q and *CDKN2A* loss whilst FISH is inadequate. Fusion detection was successful, but rare novel fusions need more extensive technologies. Finally, we propose test sets to guide future diagnostic approaches.

**Key points:** We evaluated and cross-validated ependymoma biomarkers in a large prospective clinical trial cohort.

Accurate biomarker evaluation is critical to the success of clinical trials and patient care.

We propose core and core^+^ biomarker test sets for future molecular stratification.

**Importance of the Study:** High-risk paediatric ependymoma has a poor prognosis and is devastating at relapse. Molecularly defined ependymoma types need to be accurately and reliably linked to biomarkers to predict clinical outcomes and design clinical trials. Here, we evaluated and cross-validated ependymoma biomarkers in a large prospective clinical trial cohort highlighting the importance of systematic evaluation of different methods. We provide evidence to guide test selection to support the molecular stratification of paediatric ependymoma and deliver insights into the rationalisation of biomarkers for use in resource-limited settings.

## INTRODUCTION

The management of ependymoma in children and young adults is complex and the clinico-bio-pathological correlates of outcome remain poorly understood. Overall, prognosis remains poor in most patients and at relapse is dismal^1^. Over half of patients ultimately die from the disease and survivors face significant long-term sequelae. Half of cases occur under the age of five years, a time in which the infant brain is undergoing rapid development and therefore at heightened risk of harm from medical interventions^2–7^.

Prior to the last decade, ependymomas were defined by anatomical location. However, the advent of the DNA methylation-based classification of ependymal tumours has improved our understanding by delineating multiple distinct tumour types and subtypes^2,3^. The latest World Health Organisation Classification of Tumours of the CNS (WHO CNS5)^3,8^ now defines ependymoma by both anatomical and molecular characteristics. It is critical to facilitate the identification, prognostication and stratification of ependymoma by linking these molecular tumour types to robustly validated biomarkers.

The extent of tumour resection represents the most reproducible clinical prognostic factor to date, with gross total resection (GTR) correlated with improved survival in multiple studies^1,9–14^. Despite this, many patients with GTR experience relapse, calling for validation of previously proposed biomarkers^7,15–19^ in a prospective multicentre clinical trial setting. Additionally, work is needed to understand the best way to measure the accuracy and reproducibility of these biomarkers.

An aim of the SIOP Ependymoma II clinical trial is to identify and validate prognostic biomarkers within the collaborative “Biomarkers of Ependymoma in Children and Adolescents (BIOMECA)” study^20^. In this first prospective BIOMECA study, we compare molecular pathology methods across the first 147 consecutive cases from the SIOP Ependymoma II trial across six European laboratories. We aimed to determine the most accurate and reproducible methods for the analysis of pre-defined high-priority biomarkers in a clinical trial context.

The methods evaluated include epigenetic profiling via EPIC 850K methylation arrays; immunohistochemistry (IHC) for nuclear p65-RELA, H3K27me3, and Tenascin-C (TNC); copy number analysis via FISH (fluorescent *in situ* hybridisation) and MLPA (multiplex ligation-dependent probe amplification; 1q, *CDKN2A*), and MIP (molecular inversion probe; whole genome) and DNA methylation array (whole genome); the analysis of *ZFTA-* and *YAP1*-fusions by RT-PCR, sequencing, Nanostring and break-apart FISH.

## METHODS

### Patients and clinical specimens

The first 147 consecutively enrolled cases in the SIOP Ependymoma II clinical trial (trials.gov identifier: NCT02265770) from the UK, France, Spain, Czech Republic and Ireland were included. All patients had a newly diagnosed ependymoma, confirmed by central neuropathological review according to the revised WHO 2016 classification^20^. All analyses were performed on whole sections of formalin-fixed and paraffin-embedded (FFPE) primary samples. Nottingham 2 Research Ethics Committee of the National Health Service Health Research Authority (HRA) gave ethical approval for this work (Reference: 15/EM/0103). Written consent was obtained prior to study enrolment.

### Evaluation of methods and techniques

To evaluate reproducibility, techniques were conducted across six European BIOMECA national reference laboratories (Suppl. Tbl. 1). Each marker was analysed for inter-centre concordance using Cohen’s or Fleiss’s kappa. K values >0.41 indicate moderate agreement, values >0.61 indicate substantial agreement^21^. DNA methylation profiles were used as criterion standard for tests of specificity and sensitivity. Sensitivity was calculated by measuring the ratio of true positives to all positives and specificity by measuring the ratio of true negatives to all negatives. Test accuracy was calculated by measuring the proportion of correct results compared to the criterion standard (DNA methylation profiles). p65 immunohistochemistry for the diagnosis of supratentorial ependymoma with RELA fusions was assessed against a standard criterion comprised of DNA methylation profiling and identification of fusions via PCR and/or targeted sequencing approaches. Where no methylation profiling result was available cases were excluded from sensitivity and specificity measurements.

### Fluorescent in situ hybridization (FISH; 1q25, *ZFTA-, YAP1-* fusions)

FISH for chromosome 1q gain was performed using commercial 1q25/1p36 probes on FFPE sections (4μm) according to manufacturers’ instructions in the UK and France (Suppl. Methods). FISH for *ZFTA-* and *YAP1-*fusions was performed on interphase nuclei as previously described^16^.

### DNA extraction and copy number analysis

Extracted DNA was assessed for copy number variation via multiplex ligation-dependent probe amplification assays (MLPA; chromosome 1q and *CDKN2A*; UK), molecular inversion probe assays (MIP; whole genome; Bonn) and EPIC 850K methylation array (whole genome; DKFZ) (Suppl. Methods).

### Immunohistochemistry (H3K27me3, TNC, and nuclear p65-RelA)

Whole FFPE sections (4μm) immunostained in three BIOMECA centres (Supp. Tbl. 1; Suppl. Methods). H3K27me3, TNC and nuclear p65-RelA staining was double-scored as positive or negative.

### EPIC 850K DNA Methylation array

DNA methylation array was performed in the UK BIOMECA laboratory in conjunction with UCL Genomics, London (Suppl. Methods) and at the German Cancer Research Centre (DKFZ), Heidelberg as previously described^2,18^. Array data was analysed using the Heidelberg Brain Tumor Methylation Classifier (www.molecularneuropathology.org, version 12 (V12). A classifier score of 0.9 was applied as a cut-off for confident methylation class prediction.

### RT-PCR, Sequencing and Nanostring (*ZFTA-* and *YAP-*fusion)

In the Como BIOMECA laboratory, RT-PCR was performed to detect common variants of fusions of *ZFTA-RELA* (type 1, exon 2-2; and type 2, exons 3-2), *YAP1-MAMLD1* (exons 5-3 or 6-2), *ZFTA-MAML2* (exons 5-2), and *ZFTA -YAP1* (exons 5-1) (Suppl. Methods). *ZFTA-RELA* fusion transcript was investigated by TaqMan real-time PCR. All data was analysed with Sequencing Analysis Software (Applied Biosystems).

In the Bonn BIOMECA laboratory, the presence of *ZFTA-* and *YAP1-MAMLD1* fusions was examined by RT-PCR as previously described^24,25^. Further molecular analysis of gene fusions was implemented with the Nanostring fusion panel. Four ZFTA-like classified cases were examined further with the Next-generation mRNA gene fusion panel using the TruSight Fusion Panel (Illumina, San Diego, CA, USA) as previously described^22^. Sequencing data were analysed by the Arriba tool (https://github.com/suhrig/arriba)^23^.

## RESULTS

### Case Cohort

147 tumours accrued from two national centres and three partner centres were included (Tbl. 1). There was an even gender balance (males, n=78, 53%, females n=69, 47%). Median age at diagnosis was 40 months (Range: 5-225). 76% were infratentorial (posterior fossa; PF, n=111), 22% supratentorial (ST, n=32), and 3% spinal (SP, n=4). (Suppl. Tbl. 4).

**Table 1:** Summary of case cohort and patient characteristics.

| Case cohort and patient characteristics |  |  |  |  |
| --- | --- | --- | --- | --- |
| PF = posterior fossa, ST = supratentorial, SP = spinal |  |  |  |  |
| Characteristic | Overall, N = 147 | PF, N = 111<sup>†</sup> | SP, N = 4<sup>†</sup> | ST, N = 32<sup>†</sup> |
| **Country** |  |  |  |  |
| UK | 65 (44%) | 54 (49%) | 4 (100%) | 7 (22%) |
| France | 62 (42%) | 42 (38%) | 0 (0%) | 20 (62%) |
| Spain | 16 (11%) | 12 (11%) | 0 (0%) | 4 (12%) |
| Czech Republic | 3 (2.0%) | 2 (1.8%) | 0 (0%) | 1 (3.1%) |
| Ireland | 1 (0.7%) | 1 (0.9%) | 0 (0%) | 0 (0%) |
| **Gender** |  |  |  |  |
| M | 78 (53%) | 64 (58%) | 0 (0%) | 14 (44%) |
| F | 69 (47%) | 47 (42%) | 4 (100%) | 18 (56%) |
| **Age (months)** |  |  |  |  |
| ≥36 | 80 (54%) | 51 (46%) | 4 (100%) | 25 (78%) |
| ≤36 | 67 (46%) | 60 (54%) | 0 (0%) | 7 (22%) |
| <sup>†</sup> Statistics presented: n (%) |  |  |  |  |

### Methylation profiling

Application of v12.5 of the Heidelberg Brain Tumour Methylation Classifier resulted in a calibrated score ≥0.9 in 91.1% (134/147) of all cases (PFA: 96/134, PFB: 10/134, ST-ZFTA: 22/134, ST-YAP1: 1/134, SP-MPE: 3/134, ST-PLAGL1: 2/134; Fig. 1). 11 cases (7.5%) did not reach the cut-off of 0.9. However, manual inspection of the t-SNE showed that 7/11 of these samples clustered within (1/11) or close to (6/11) the cluster of their best prediction (4/11: PFA, 2/11: ST-ZFTA, 1/11: ST-YAP1) (Suppl. Fig. 1). In 2 cases (1.4%) no score was generated.

**Figure 1.**
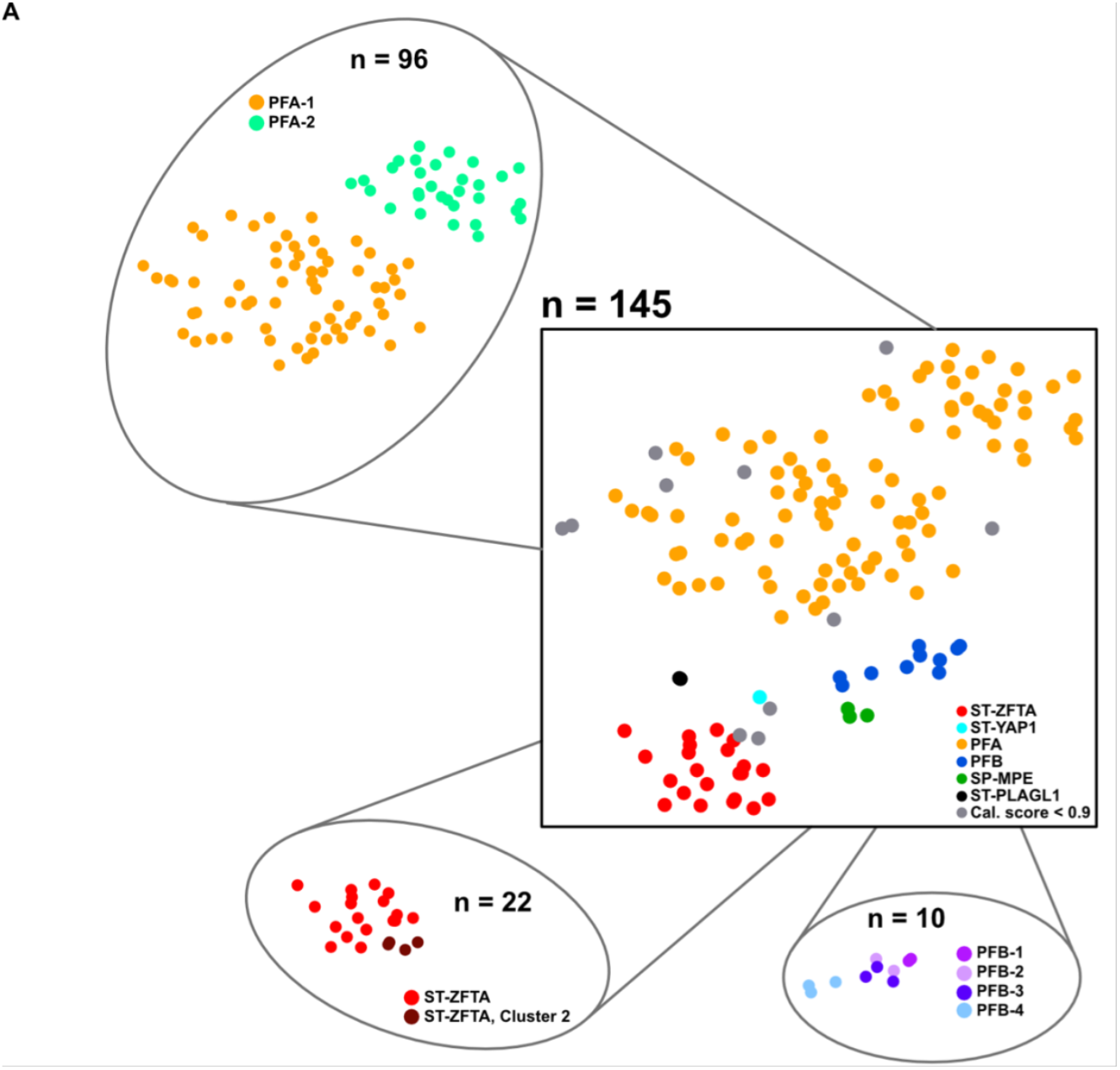
DNA-methylation profiling results. t-SNE plot visualizing DNA-methylation based clustering of the BIOMECA cohort. Central t-SNE: Molecular groups, satellite t-SNEs: molecular subgroup for PFA, molecular subtype for ST-ZFTA and PFB. Samples are colorized according to the best available prediction.

PFA can be further stratified into two main subgroups (PFA-1/2) and nine subtypes (1a – f and 2a – c) ^24^. All 96 PFA cases also had a score ≥ 0.9 for one of the two PFA subgroups, with 67.8% (65/96) PFA-1, and 32.2% (31/96) PFA-2. These frequencies reflect those in the original study describing PFA subclassification (Fig. 1)^24^. PFB subtyping resulted in confident prediction scores for 10/10 cases (PFB1: 2/10, PFB2: 2/10, PFB3: 3/10, PFB4: 3/10) (Fig. 1)^25^. Recently, we have described further ST-ZFTA heterogeneity, with additional subgroups characterized by various histological appearances and alternative *ZFTA*-fusions^5^. Out of the 22 patients predicted as ST-ZFTA, 18 had calibrated scores ≥0.9 for classic ST-ZFTA, which normally harbor *ZFTA-RELA* fusions, whilst four were stratified into the alternative ZFTA-like subtype 2 (Fig. 1).

### Evaluation of methods to assess copy number variation

#### DNA Methylation Assays

Methylation array-derived Copy Number Variation (CNV) plots were analysed with a focus on previously described copy number alterations within the respective molecularly defined types (Fig. 2)^2,5,24–26^. A gain of chr. 1q was present in 4/22 ST-ZFTA, 14/96 PFA and 1/10 PFB, respectively (Fig. 2; Suppl. Tbl. 4). *CDKN2A* loss and chromothripsis on chr. 11 were restricted to ST-ZFTA, while chr. 22 loss was present in ST-ZFTA (6/22), PFB (7/10) and PFA (4/96), as previously described (Fig. 2; Suppl. Tbl. 4)^2,5,24,25^. As previously described ^24^, chr. 1q-gains were enriched in PFA1c, representing a particularly aggressive form of PFA (Suppl. Fig. 1B; Suppl. Tbl. 6).

**Figure 2.**
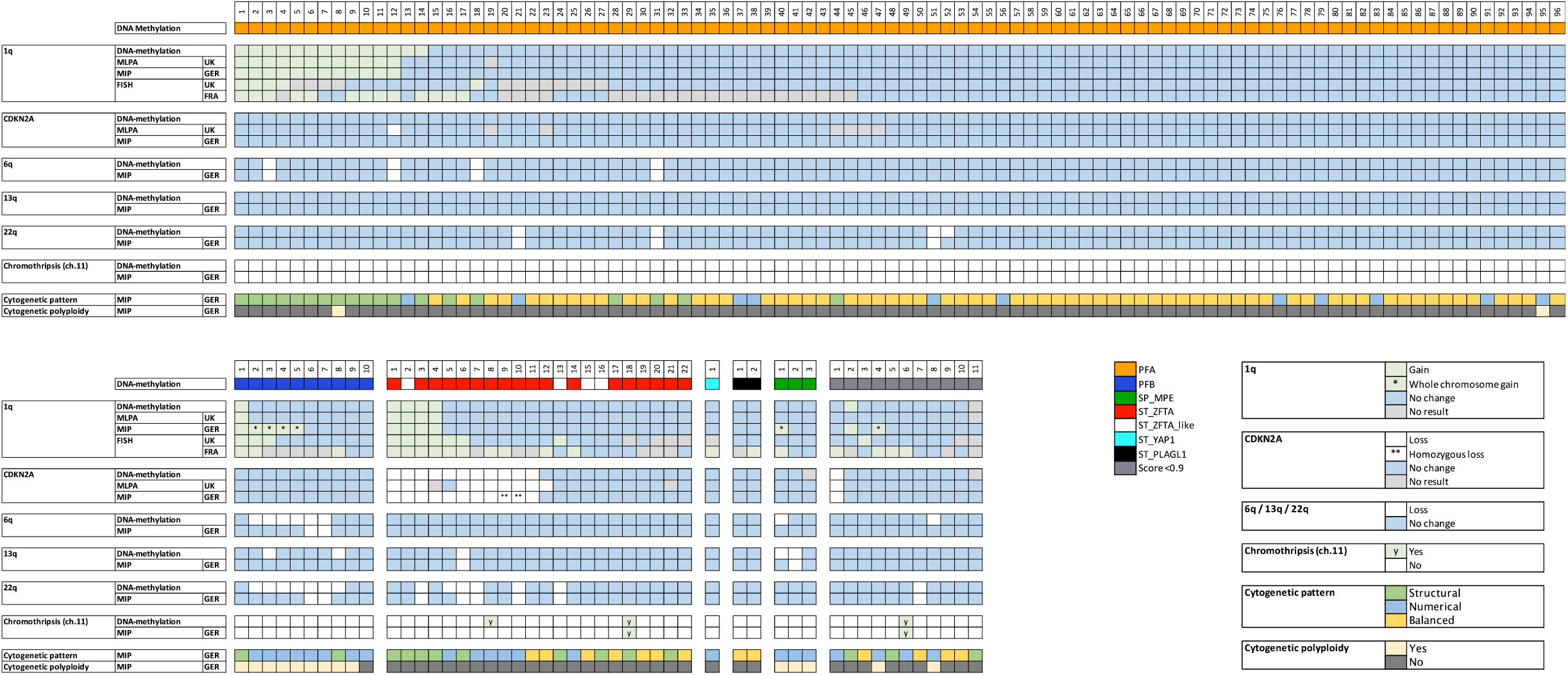
Evaluation of methods to assess copy number alterations.

#### Molecular Inversion Probe (MIP) Assays

High-resolution, quantitative MIP arrays revealed chr. 1q gain in 12/96 PFA, 1/10 PFB and 4/22 ST-ZFTA (Fig. 2). MIP analysis identified 4/10 PFB and 1/3 SP-MPE as exhibiting whole chr.1 gain. Loss of *CDKN2A* and 13q, and chromothripsis at chr. 11, was not observed in PFA or PFB using MIP. Chromothripsis at chr.11 was observed in one ST-ZFTA, whilst loss of 13q was observed in one ST-ZFTA and one SP-MPE (Fig. 2). *CDKN2A* loss was detected exclusively in 11 ST-ZFTA, with homozygous loss observed in 2/11 (#9-10, Fig. 2; Suppl. Fig. 2). Loss of 6q was observed in 4/96 PFA and 2/10 PFB. Co-occurrence of 1q/6q was documented in one case (PFA, #12, Fig. 2; Suppl. Fig. 3). Loss at chr.22 was detected in 3/96 PFA, 2/10 PFB, and five ST-ZFTA.

MIP assays provided information regarding cytogenetic alterations/pattern and ploidy. A polyploid cytogenetic pattern was observed in 9/10 PFB and 3/3 SP-MPE (Fig. 2) and demonstrated mostly numerical alterations (8/10 PFB). PFAs revealed 66 balanced, 19 structural and 11 numerical cytogenetic alterations (n=96; Fig. 2). In contrast, ST-ZFTA showed a more equal mix of 7 balanced, 9 structural and 6 numerical cytogenetic alterations (n=22; Fig. 2).

For 1q gain, MIP assays demonstrated sensitivity and specificity of 94.4% and 99.1% respectively. Test accuracy was 98.4% compared with methylation-based assessments (Tbl. 2).

**Table 2:**
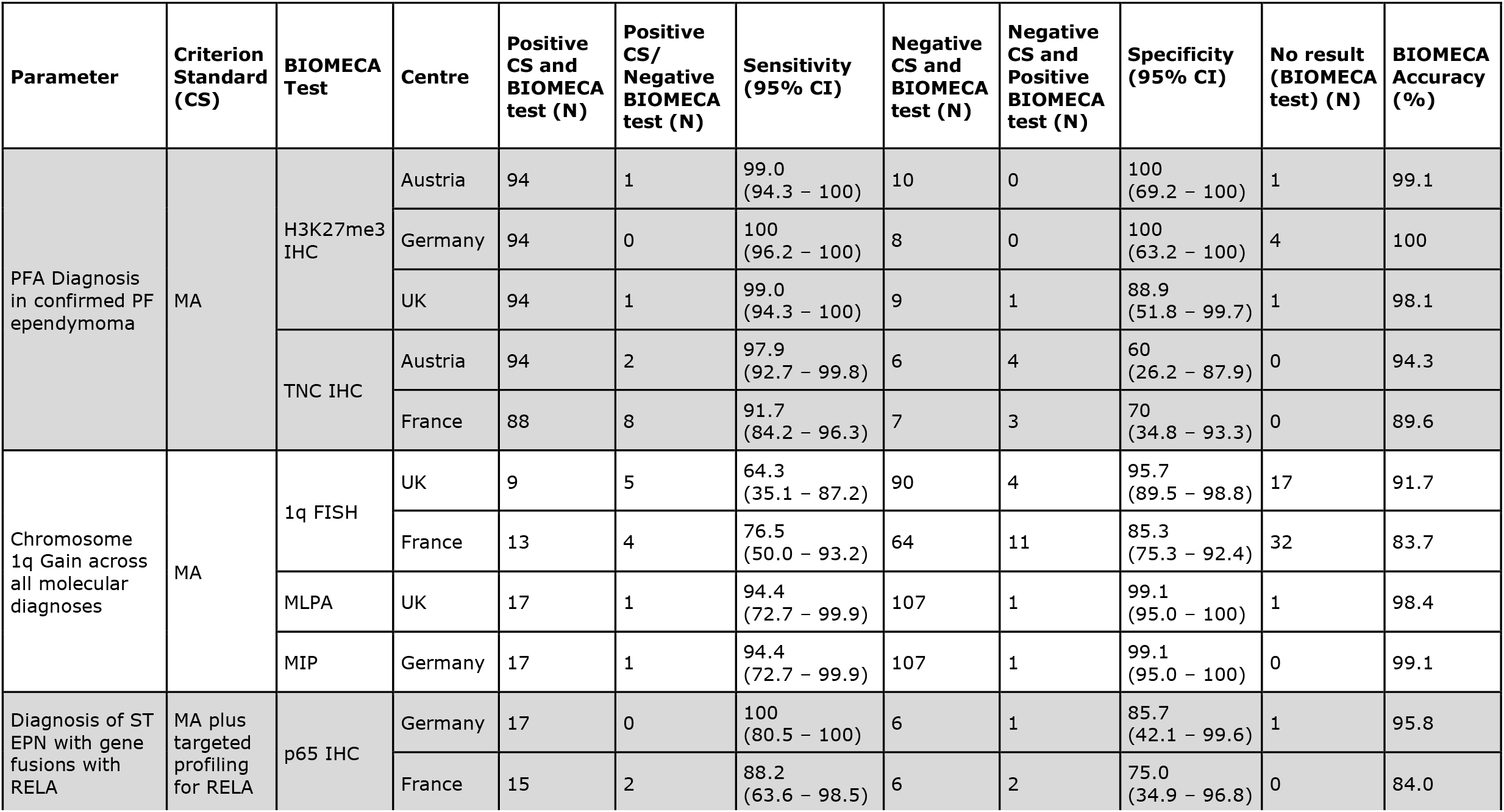
Summary of accuracy for key BIOMECA tests under evaluation stratified by centre and test type. BIOMECA tests with equivocal or no result removed from calculation. For gain of chromosome 1q only partial chromosomal gains included in analysis – cases with whole chromosome excluded as known to only be detectable with MIP. Test accuracy calculated by true positives and true negatives divided by all test results. Confidence intervals calculated via the exact binomial approach. CS: Criterion Standard, MA: DNA Methylation Array, PF: Posterior Fossa, ST: Supratentorial, IHC: Immunohistochemistry, FISH: Fluorescent In-Situ Hybridisation, CI: Confidence Interval.

#### Multiplex-Ligation Dependent Probe Amplification (MLPA) Assay

Chr. 1q gain and *CDKN2A* loss were assessed using MLPA. After adjusting the analysis for whole chr.1 gains identified via MIP all three DNA-based techniques (MIP, methylation array and MLPA) demonstrated 98.4% concordance for assessing chr. 1q gain (n=134; Fleiss’ k=0.958, p=0) and 90.3% concordance for *CDKN2A* (n=134; Fleiss’ k=0.655, p=0). MLPA did not yield a CDKN2A result in eight cases. MIP and DNA methylation profiles for *CDKN2A* demonstrated 98.5% concordance (n=134; Cohen’s k=0.91, p<0.0001).

MLPA had sensitivity and specificity of 94.4% and 99.1% respectively for 1q gain. Test accuracy was 98.4% compared with methylation-based assessments (Tbl. 2), identical to those for MIP.

#### Fluorescent In-Situ Hybridisation (FISH)

Chr. 1q FISH demonstrated low inter-centre concordance (n=134; 57.5%, Cohen’s k=0.152, p=0.0191). For cases classified using DNA methylation array, gain at chr. 1q was reported as 11.2% (15/134) and 21.6% (29/134) in the UK and France respectively. However, concordant observation was only reported in 8.2% (11/134; 3 PFA, 2 PFB, 6 ST-ZFTA; Fig. 2). As similarly reported by Andreiuolo *et al*.^27^, a significant number of cases, 26% (35/134, France) and 12.7% (17/134, UK) showed technical failure. The centre in France determined 16.4% (22/134) UK cases to have failed compared to just 5.2% (7/134) UK cases determined to have failed in the UK centre. This may reflect differences in tissue processing protocols in respective centres.

In addition to the discordant results and technical failures experienced via FISH, measures of accuracy were also poor. In the UK, FISH for 1q gain was associated with a sensitivity and specificity of 64.3% and 95.7% respectively, whilst the same measures in France were 76.5% and 85.3%. Accuracy was 91.7% in the UK and 83.7% in France (Tbl. 2).

### Evaluation of H3K27me3 and Tenascin-C immunohistochemistry

In all PF cases with a classifier calibration score ≥ 0.9 H3K27me3 and TNC expression were assessed via IHC to investigate utility as a surrogate marker for PFA/PFB.

H3K27me3 expression demonstrated 92.5% inter-centre concordance with agreement in 94.8% (91/96) PFA and 60% (6/10) PFB cases (n=106; Fleiss’ k=0.732, p<0.000; Fig. 3). Specifically, PFA demonstrated a loss of H3K27me3 expression, whilst PFB cases retained expression. Across three centres, loss of H3K27me3 had a specificity of 99%, 100% and 99% and specificity of 100%, 100% and 88.9% for diagnosing PFA in PF ependymoma. Test accuracy ranged from 98.1% - 100% (Tbl. 2). Of 11 cases of PF ependymoma with classifier score <0.9, H3K27me3 staining identified 63.6% (7/11) as PFA. Seven of these 11 cases clustered close to the clusters of their respective best prediction, demonstrating that visual inspection of t-SNE or other dimensional reduction visualisations can be useful in cases with ambiguous classification scores.

**Figure 3.**
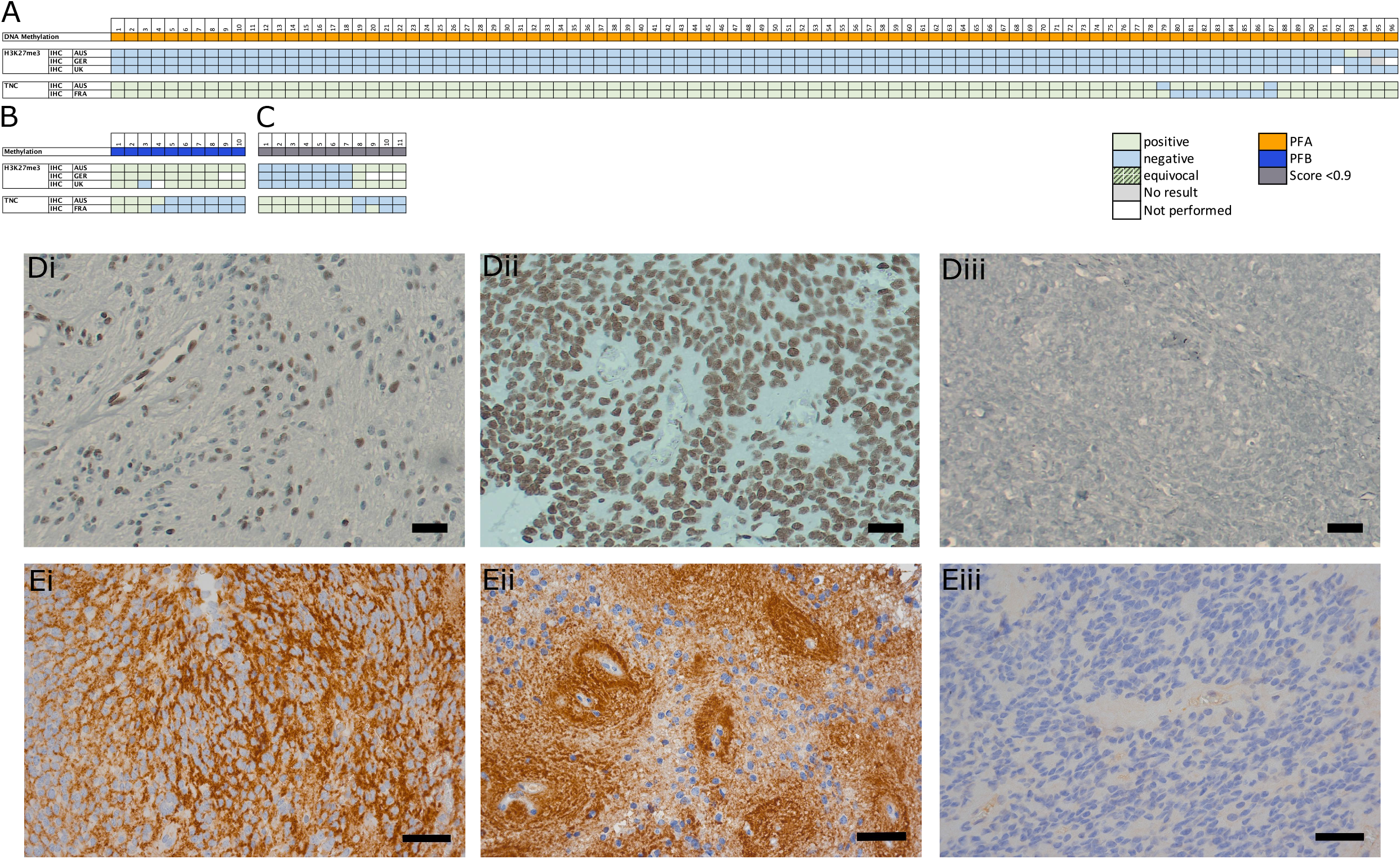
Evaluation of IHC for H3K27me3 and TNC as potential surrogate PFA markers. Posterior fossa tumours classed as PFA (A) and PFB (B) via DNA-methylation array (calibration score ≥0.9) and the immunohistological result per case for H3K27me3 and TNC IHC as assessed per centre. (C) H3K27me3 and TNC results in cases not classified by DNA methylation array. (D) IHC images representing H3K27me3 in a PFA (i) and a PFB (ii) case. (E) IHC images representing TNC pericellular (i) and perivascular (ii) expression. (D-E) Original magnification X40, scale bars 50mm. Image of representative negative controls for H3K27me3 (Diii) and TNC (Eiii).

TNC expression demonstrated a 91.5% inter-centre concordance between the two centres performing the analysis (n=106; Cohen’s k=0.561, p<0.000; Fig. 3). Concordant positive TNC staining was observed in 89.6% (86/96) of PFA and only 30% (3/10) of PFB. 60% (6/10) of PFB demonstrated concordant negative staining for TNC, whilst one case was discordant between centres. Positive staining for TNC as a predictor of PFA diagnosis in PF ependymoma had sensitivities of 97.9% and 91.7% and specificities of 60% and 70%. Test accuracy was 94.3% and 89.6%.

Assessing these markers together, concordant loss of H3K27me3 and simultaneous expression of TNC was observed in 86.5% (83/96) of PFA (Figure 3C).

### Assessment of methods used for the detection of *ZFTA-* and *YAP1*-fused ependymomas

The detection of molecularly defined *ZFTA-* and *YAP-*fused ependymomas was assessed using IHC, FISH, RT-PCR, sequencing and Nanostring technology.

IHC for nuclear p65-RelA protein was assessed on 23 *ZFTA-* and *YAP1-*fused tumors, repeated across two centres (Fig. 4A). ST-ZFTA demonstrated an 86.4% (18/22) inter-centre concordance (n=22; Cohen’s k=0.582, p=0.0058; Fig. 4A). 68.2% (15/22) of these cases demonstrated concordant positive staining for nuclear p65-RelA protein and 3/22 cases, concordant negative staining. Whilst only one ST-YAP1 case (case #25; Fig. 4A) was identified in this cohort, this was negative for nuclear p65-RelA in both centres.

**Figure 4.**
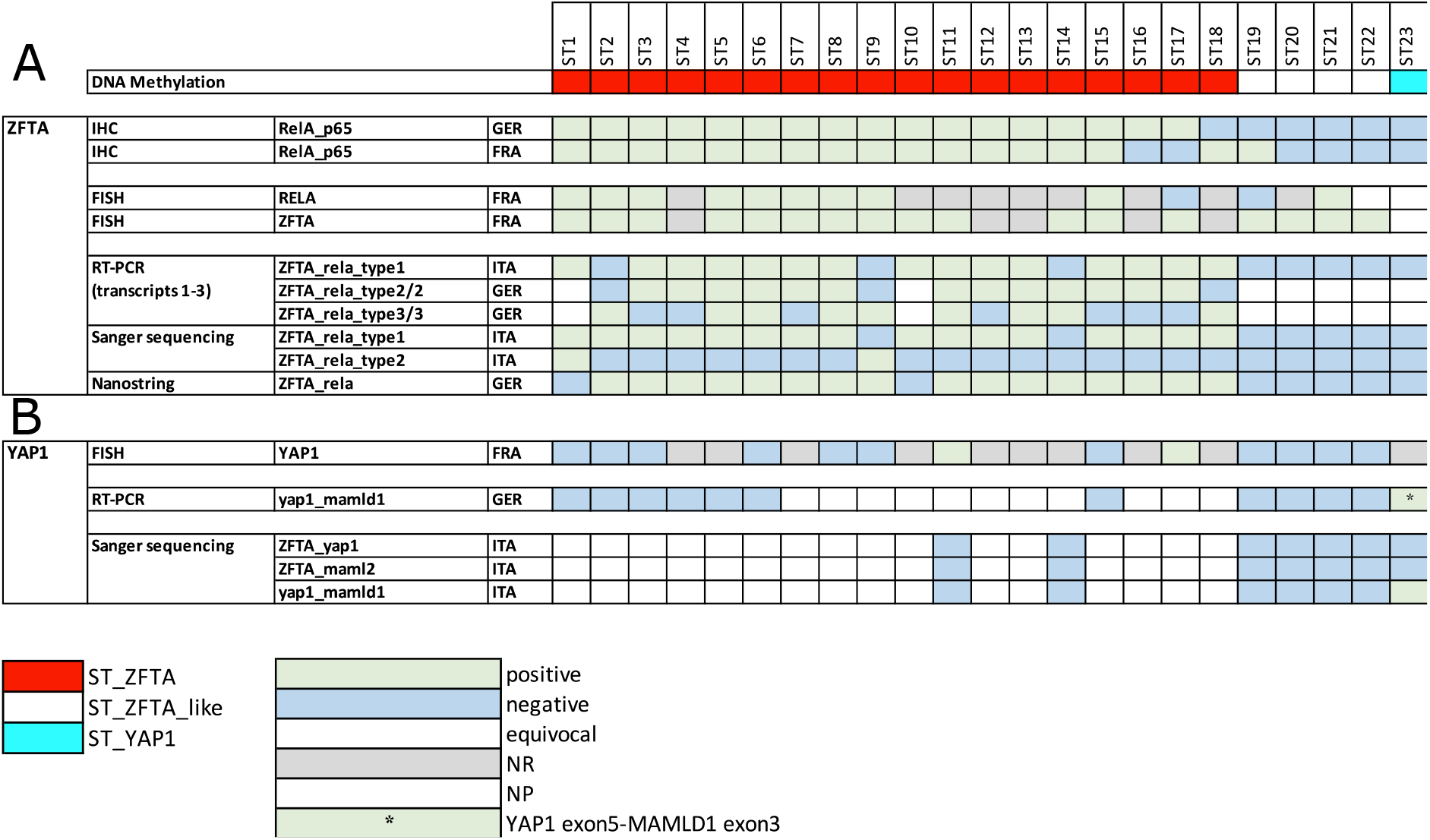
Comparison of methods used to assess *ZFTA*-(A) and *YAP1*-(B) fusions in supratentorial tumours.

FISH revealed 45.5% (10/22) ST-ZFTA cases with rearrangements at both the *RELA* and *ZFTA* loci (Fig. 4A). One case gave an equivocal result, and two showed a rearrangement at the *ZFTA*, but not the *RELA*, locus. FISH failed in nine cases assessed for rearrangement at the *RELA* locus, and five cases at the *ZFTA* locus. 41% (9/22) ST-ZFTA cases demonstrated concordance of positive nuclear p65-RelA IHC staining with simultaneous rearrangements at both the *RELA* and *ZFTA* loci via FISH.

RT-PCR with subsequent Sanger sequencing and Nanostring assay were used to investigate and confirm the presence of *ZFTA-RELA* fusions. Fusion transcripts were detected in 78.3% (18/22) molecularly defined cases in 2 centres conducting the analysis. *ZFTA-RELA* transcripts 1-3 were detected in all those classified as classic ST-ZFTA (cases #1-18, Fig. 4A) via DNA methylation array, with no such fusion transcripts detected in the four cases classified as the alternative ZFTA-like subgroup (cases #19-22, Fig. 4A). IHC for nuclear p65-RelA in the ZFTA-like cases was concordantly negative in 3/4 cases in two centres. Interestingly, in all four ZFTA-like cases, a rearrangement at the *ZFTA* locus was observed via FISH analysis, whilst rearrangement at the *RELA* locus was observed in one case, plus one equivocal result. RNA sequencing of these four ZFTA-like cases identified fusions of *ZFTA-NCOA2* in case #19, *ZFTA-NCOA1* in case #20 and *ZFTA-MAML2* in cases #21 and #22 (Suppl. Fig. 3)

Case #23 (Fig. 4), classified as a YAP1 tumour, demonstrated an equivocal result via FISH analysis for rearrangement at both the *ZFTA* and *RELA* locus, but was not positive for nuclear p65-RelA IHC and was negative for all *ZFTA-RELA* transcripts via RT-PCR, Sanger sequencing and Nanostring assay. FISH analysis for YAP1 failed for this case. However, RT-PCR detected a *YAP1-MAMLD1* fusion (*YAP1-exons 5/MAMLD1-exon3*), which was confirmed by Sanger sequencing (Fig. 4B).

When a combination of DNA methylation profile and fusion transcript analysis by targeted sequencing or PCR were combined as the criterion standard, p65 IHC had sensitivities of 100% and 88.2% and specificities of 85.7% and 75% in identifying supratentorial fusions which contained *RELA* as the partner with *ZFTA* (Tbl. 2).

## DISCUSSION

Our study aimed to establish the most accurate and reproducible techniques for measuring key ependymoma biomarkers across 147 consecutive samples from SIOP Ependymoma II trial participants enrolled in the pan-European “Biomarkers of Ependymoma in Children and Adolescents (BIOMECA)” study. BIOMECA is the first pan-European study that has evaluated and cross-validated ependymoma biomarkers in a large prospective clinical trial cohort.

We were able to show that H3K27me3 IHC is both accurate and reproducible for the diagnosis of PFA ependymoma in a clinical trial setting. Additionally, DNA methylation profiling, MIP and MLPA are all effective techniques for assessing key copy number changes in this disease. Combinations of IHC, PCR and targeted sequencing are suitable for the delineation of fusion gene status in supratentorial ependymomas. Our study suggests that TNC is not a useful marker for PF ependymoma, and that FISH should be abandoned as a technique for the assessment of copy number status in ependymoma.

The integration of tumour-specific histopathology and molecular profiling is gaining pace, the updated 2021 WHO CNS5 classification of CNS tumours now lists ten molecularly defined types of ependymal tumours^3^. Understanding the role of prognostic biomarkers for each of these entities is essential for the evolution of precision medicine and tailored therapy for ependymoma, and to understand how markers can be rationalised for use in both clinical trials and limited resource settings.

All cases included in this study were diagnosed according to local neuropathology review before confirmation by central review and DNA methylation profiling^2,3,28,29^. DNA methylation profiling resulted in a confident diagnosis of a molecularly defined type for 91.1% of all patients. Only 2/147 cases could not be profiled due to poor-quality DNA from FFPE tissue highlighting DNA methylation profiling as the criterion-standard for this assessment. The importance of DNA methylation profiling was also highlighted by the two supratentorial tumours diagnosed as ependymoma, which clustered with neuroepithelial tumours with *PLAGL1* fusions^30^, demonstrating that the classifier is a tool under continuous development.

A global reduction of H3K27me3 expression in EPN-PFA is used as a surrogate marker for this molecular group^31–33^ and is now recommended as an essential diagnostic criterion in the updated 2021 WHO CNS5 classification^3^. Our data aligns strongly with this recommendation. 95.7% of PFA cases demonstrated concordant loss of H3K27me3 expression across three centres. The reproducibly high sensitivity and specificity across the three centres also robustly supports the use of this marker for the diagnosis of PFA tumours. H3K27me3 expression represents a useful biomarker for settings with limited resources or where methylation profiling is not possible. In contrast, previous suggestions that TNC expression is a surrogate marker for PFA ependymomas^34–36^ is not confirmed by our data. TNC expression was found in a substantial fraction of PFB tumours and was associated with low specificity.

*RELA* encodes the p65-RelA protein which shows nuclear accumulation upon pathological activation of the NFkB signalling pathway^18,26^. Other studies have investigated the potential of IHC to predict *ZFTA-RELA* fusion status in comparison to using RT-PCR and Sanger sequencing or Nanostring^37,38^. Detection of the fusion is an essential diagnostic criterion as per the 2021 WHO CNS5 classification of supratentorial *ZFTA*-fused ependymomas, and p65-RelA IHC is now listed as a desirable marker as part of the diagnostic pathway^3^. Our data demonstrate significant inter-centre concordance using IHC and corresponding directly to those cases where fusion transcripts were detected using RT-PCR and sequencing or Nanostring, and additionally classified as ST-ZFTA using DNA methylation array. We demonstrated sensitivities of 88.2 and 100% for the identification of fusions with *RELA* as a partner to ZFTA using p65 IHC, although confidence intervals are wide in view of the low number of these cases. Where atypical fusions with partners other than *RELA* are present, p65-RelA IHC cannot help in predicting molecular class. However, as in most cases *ZFTA* is fused to *RELA*, the detection of nuclear accumulation of p65-RelA by IHC represents an easy, cost-effective, and reliable surrogate marker for most ST-ZFTA cases ^39^.

FISH has long been established in most diagnostic pathology laboratories around the world for the assessment of genomic rearrangements^40^. Here we found that the break-apart FISH technique failed to detect *RELA* and *ZFTA* fusions in 9/22 (41%) cases classed as ST-ZFTA. This may be a consequence of both *RELA* and *ZFTA* being located only 1.9Mbp apart on chromosome band 11q13^18^, making the interpretation and subsequent analysis difficult. This failure rate is higher than that observed by Pages *et al*. (^38^), where approximately 10% of supratentorial cases did not yield a result using break-apart FISH. Similarly, although only one *YAP1*-fused ependymoma case was identified in this study, FISH failed to detect the fusion. We do not recommend FISH as a primary approach for classifying supratentorial ependymomas.

Chromosome 1q gain and *CDKN2A* loss have been proposed as independent markers of worse prognosis in paediatric PFA and ST-ZFTA ependymoma respectively^7,12,15,34,41–43^. Traditionally, FISH has been the primary technique to assess chromosome 1q gain however, the optimal method for detection of 1q gain has been debated, with reports that 20% of cases cannot be assessed by FISH on FFPE tissue^34^. This study does not support FISH as a reliable method for assessment of 1q gain owing to the high failure rate observed by two centres (13-26%) and low (57%) inter-centre concordance. Additionally, the sensitivity associated with FISH in detecting 1q gain when compared to methylation-based techniques was just 64.3% and 76.5%. Whilst FISH protocols may be optimised and standardised within a centre, variation of tissue processing between centres may significantly impact systematic biomarker assessments. This cannot be avoided in a study where tissue samples are collected in multiple international centres, explaining some of the discordance observed in this study.

In assessing chromosome 1q gain and *CDKN2A* loss, we compared copy number analysis via three molecular methodologies: DNA methylation array, MIP and MLPA. Concordance for the identification of gain at chromosome 1q (98.4%) and loss of *CDKN2A* (90.3%) using these three methods was high. Furthermore, whole genome-wide copy number analysis was enabled with DNA methylation array and MIP. Sensitivity, specificity, and accuracy for both MIP and MLPA were comparably high when compared with DNA methylation profiling and based on this measure alone all three techniques are appropriate for the identification of 1q gain in PFA ependymoma. However, the high-resolution, genome-wide MIP technology revealed quantitative copy-number information in all tumours enrolled in this study. This technology works with DNA input down to 20ng in contrast to DNA methylation profiling which needs significantly more. Additionally, in contrast to MIP, DNA methylation-based CN calls cannot be adjusted for diploidy. This adjustment, however, is mandatory for exact copy number calls, particularly in the complex polyploid genomes occurring in PFB and MPE. DNA methylation array analysis was unable to identify some chromosomal losses in complex genomes such as PFB but, critically, was able to reliably detect gains of chromosome 1q in PFA tumours. Similarly, the distinction between hemizygous and homozygous CDKN2A deletion is more secure after diploid correction, however, CDKN2A deletions were correctly identified in all samples using DNA methylation. The ability of DNA methylation profiling to reliably detect 1q gain and CDKN2A deletions, alongside its wider availability, makes it the preferred tool for molecular stratification in urgently needed clinical trials in poor outcome ependymoma subgroups. MLPA is a similar DNA-based method as MIP, however it scores only around 20 probes compared to high-resolution MIP with more than 300,000 probes distributed over the genome. In this study, no technical failures were reported for MIP compared to a small number with MLPA (6.1%) and DNA methylation array (2%).

From our data we propose the concept of applying techniques in a CORE and CORE^+^ model, which aligns with the 2021 WHO CNS5 classification’s essential and desirable criteria for ependymoma diagnosis. CORE tests represent those that can currently be used to stratify and inform clinical trials and diagnosis and include immunohistochemistry and DNA methylation profiling. CORE^+^ tests have additional advantages for challenging cases and for use in the research setting and comprise of MIP and RNA-NGS sequencing.

All ependymoma subgroups can be profiled using CORE techniques. Using IHC initially as recommended enables a cost-effective, well-established and available technique. IHC can be reliably used as a surrogate means to detect *ZFTA-RELA-*fused (nuclear p65-RelA) and PFA (loss of H3K27me3) ependymomas. DNA methylation profiling represents a powerful tool for classification of ependymoma where histopathological features may converge on more than one possible molecularly defined tumour type, examples include tumours with *BCOR* internal tandem duplication (ITD), astroblastomas with *MN1* alteration or tumours with *PLAGL1* fusions^5,30,44^. However, it is recognised that access to DNA methylation arrays varies, especially in low and middle-income countries, so continued development of techniques not based on complex molecular methodologies to confidently classify brain tumours is vital.

Whilst CN information, particularly the important ependymoma biomarkers 1q gain and CDKN2A, can be reliably obtained with DNA methylation array, in cases where there are complex cytogenetic patterns or paucity of tissue, high-resolution, quantitative MIP arrays can be utilised. However, access is currently less widely available to centres that may participate in clinical trials of the future. Therefore, whilst MIP is not part of our core set recommendations, in the instances outlined above it can be used as a non-mandatory CORE^+^ assessment. Similarly, if rare fusion events must be detected in supratentorial ependymomas, RNA-NGS sequencing should also be included as Core^+^ assessment.

Biological systems are complex and multidimensional. Measuring multiple biomarkers and taking a variability-reductionist approach to interpreting outcomes will provide better information for future treatment stratification. Considering the relative rarity of ependymoma, it is of paramount importance that future prospective trials utilise a standardised and reliable set of diagnostic and prognostic markers. The BIOMECA study makes recommendations for standardising ependymoma biomarkers across clinical trials in the years to come.

## Data Availability

All data produced in the present study are available upon reasonable request to the authors

## Acknowledgements

We thank clinical research teams from contributing CCLG centres and Tissue Bank for sample provision and clinical information, and the Birmingham CRUK clinical trials unit for study support. We thank Monika Mauermann for technical assistance. We thank the Microarray Unit of the Genomics & Proteomics Core Facility, German Cancer Research Center (DKFZ) and University College London (UCL) Genomics, London, for providing excellent services regarding methylation arrays.

