## Supplemental Methods for "Optimising biomarkers for accurate ependymoma diagnosis, prognostication and stratification within International Clinical Trials: A BIOMECA study"

**Evaluation of methods and techniques**

Analyses were conducted across six European BIOMECA national reference laboratories to evaluate reproducibility of results (Suppl. Tbl. 1). Each marker was analysed for concordance of outcome between centres using Cohen’s or Fleiss’s kappa. K values >0.41 indicate moderate agreement, values >0.61 indicate substantial agreement^1^. DNA methylation profiles were used as criterion standard for tests of specificity and sensitivity. Test sensitivity was calculated by measuring the ratio of true positives to all positive results and specificity by measuring the ratio of true negatives to all negative results. Test accuracy was calculated by measuring the proportion of correct results. DNA methylation profiles were used as the criterion standard for these calculations. Test accuracy was calculated in this way for H3K27me3 and Tenascin C immunohistochemistry in the diagnosis of PFA ependymoma and for FISH, MIP and MLPA in the diagnosis of gain of copy number of chromosome 1q. p65 immunohistochemistry for the diagnosis of supratentorial ependymoma with RELA fusions was assessed against a standard criterion comprised of DNA methylation profiling and identification of fusions via PCR and/or targeted sequencing approaches. Where no methylation profiling result was available cases were excluded from the measurements of sensitivity and specificity.

**Fluorescent in situ hybridization (FISH; 1q25, *ZFTA-*, *YAP1-*)**

FISH assessment for chromosome 1q gain was carried out using commercial 1q25 probes and 1p36 reference probes on FFPE sections (4μm) according to manufacturers’ instructions in the UK and France (UK: Vysis LSI 1p36/LSI 1q25 FISH probe (Abbott Molecular); France: ZytoLight® SPEC 1p36/1q25 Dual Color Probe kit (ZytoVision, Bremerhaven, Germany)). A minimum of 100 intact, non-overlapping nuclei were analysed per tumour sample. Gain was defined by at least 15% of the counted nuclei across the whole section where the ratio of green (1q25) to red (1p36) signals was greater than one for the green signal.

FISH for *ZFTA-* and *YAP1-* fusions was performed on interphase nuclei as previously described^2^. *ZFTA* FISH was performed using a break apart custom SureFISH probe and hybridized according to the manufacturer's recommendations for SureFISH probes (Agilent Technologies, Santa Clara, CA). *ZFTA* (Empire Genomics, Buffalo, NY) and *YAP1* break-apart probe (Empire Genomics, Buffalo, NY) were also used. Hybridizations were considered non-informative if the FISH signals were either lacking or too weak to be interpreted. Signals were scored in at least 100 non-overlapping interphase nuclei. A case was considered positive when the scored nuclei displayed a break-apart signal in at least 20% of the counted nuclei.

**DNA extraction and copy number analysis**

Copy number was assessed via multiplex ligation-dependent probe amplification assays (MLPA; chromosome 1q and *CDKN2A*; UK), molecular inversion probe assays (MIP; whole genome; Bonn) and EPIC 850K methylation array (whole genome; DKFZ). Haematoxylin-eosin (H&E) – stained sections of each case were reviewed carefully prior to DNA extraction for sufficient tumour content.

For MLPA analysis, DNA/RNA was extracted from FFPE samples using the AllPrep FFPE DNA/RNA extraction kit (Qiagen, Germany) according to manufacturer’s instructions and concentration was measured by QUBIT fluorimeter (ThermoFisher, USA). A commercially available Salsa MLPA assay for 1q25 (P216-B1/303; MRC-Holland, Amsterdam, The Netherlands) and *CDKN2A* (ME024 B2; MRC-Holland) were used and performed according to the manufacturer's protocol (MRC Holland). Controls included no template controls and positive samples with known gain of chromosome 1q. Data analysis was performed using the Coffalyser.Net software (MRC-Holland).

For MIP analysis, DNA was extracted from all cases using the QIAmp DNA Mini Tissue Kit (Qiagen GmbH, Düsseldorf, Germany) according to the manufacturer’s protocol. Genomic copy number losses and gains were accessed through a molecular inversion probe array (MIP) (OncoScan CNV Plus Array, Affymetrix, Santa Clara, CA, USA). The MIP array includes 335,000 inversion probes (version V2.0) with a median probe spacing of 2.4 kb. MIP analysis was performed using at least 80 ng tumor DNA as previously described^17^. The raw MIP data were analysed using the Nexus Copy Number 8.0 Discovery Edition software (BioDiscovery, El Segundo, CA, USA). To make copy number and loss of heterozygosity estimations we used the manufacturer’s SNP-FASST2 segmentation algorithm.

For assessment of copy number variation using methylation array, plots were derived with the conumee package as previously described^23^ and analysed independently by both DRG and KWP. Ambiguous results were discussed amongst a wider group of authors to reach consensus.

**Immunohistochemistry (H3K27me3, TNC, and nuclear p65-RelA)**

Whole FFPE sections (4μm) were distributed and immunostained in three BIOMECA centres (Supp. Tbl. 1). Where the Ventana Discovery XT automated system was not used, tissue sections were incubated at 60^o^C overnight before being deparaffinised with xylene and rehydrated through decreasing concentrations of ethanol (100 to 70%) prior to primary antibody application. Subsequently, sections were incubated with normal goat serum (Vector Labs) and followed by an endogenous peroxidase block (DAKO). Target antigens were detected using the Envision Detection Kit (DAKO) with diaminobenzidine chromogen for visualization. Antibodies used within each centre: H3K27me3 – UK/Bonn (Cell Signalling Technology #9733) and Vienna (Ventana System #07-449); TNC – Vienna (Santa Cruz, sc-25328) and CHSA (CliniSciences, Mob231-01); p65-RelA – CHSA (Cell Signalling Technology #8242) and Bonn (as previously described^3^). Sections were counterstained with Gills 3 haematoxylin (TCS Biosciences) before being dehydrated and mounted with DPX. H3K27me3, TNC and nuclear p65-RelA was double-scored as positive or negative.

**EPIC 850K DNA Methylation array**

In the UK BIOMECA laboratory, DNA samples were bisulphite converted (EZ DNA Methylation kit/DNA clean and concentrator kit; Zymo Research) and FFPE restored in-house (InfiniumHD FFPE Restore Kit; Illumina). Samples were then run on the Infinium MethylationEPIC (850K) Beadchip arrays at UCL Genomics, London.  In the BIOMECA laboratory in Germany, all samples were processed and run at the German Cancer Research Centre (DKFZ), Heidelberg as previously described^3,4^. In order to ensure high comparability and to rule out technical batch effects, ten samples were profiled independently at both centres. These showed high consistency in molecular class prediction (Suppl. Tbl 2). Array data was analysed using the Heidelberg Brain Tumor Methylation Classifier ([www.molecularneuropathology.org](http://www.molecularneuropathology.org), version 12 (V12). Following the fifth edition of the WHO classification of CNS tumours^5^ and to account for the increased complexity of molecular ependymoma classification, three scores were calculated for each case: molecular group, subgroup, and subtype, respectively (Suppl. Tbl. 3). A score of 0.9 was applied as a cut-off for confident methylation class prediction.

**RT-PCR, Sequencing and Nanostring (*ZFTA-* and *YAP-*fusion)**

In the BIOMECA reference laboratory in Como, RNA was extracted from FFPE tumour samples using Maxwell16 LEV RNA FFPE Kit with Maxwell16 Instrument (Promega Corporation, Madison, WI, USA), quantified spectrophotometrically, and 500-1000ng was reverse-transcribed using Superscript III reverse-transcriptase (Invitrogen). PCR was performed with 50-100 ng input cDNA and AmpliTaq Gold DNA Polymerase (Applied Biosystems) using primers to detect the most common variants of fusions of *ZFTA-RELA* (type 1, exon 2-2; and type 2, exons 3-2), *YAP1-MAMLD1* (exons 5-3 or 6-2), *ZFTA-MAML2* (exons 5-2), and *ZFTA -YAP1* (exons 5-1). Primer sequences were as previously reported (Parker et al. 2014^6^). cDNA synthesis quality control was performed by PCR amplification of housekeeping gene PGK1 using primers 5’-CAGTTTGGAGCTCCTGGAAG-3’ (forward) and 5’-TGCAAATCCAGGGTGCAGTG-3’ (reverse).

PCR products were purified with Exosap-IT PCR Product Cleanup (Applied Biosystems) and sequencing reactions were performed using BigDye Terminator v1.1 Cycle Sequencing kit (Applied Biosystems). Electropherograms were obtained by capillary electrophoresis on SeqStudio Genetic Analyzer (Applied Biosystems) and analysed with Sequencing Analysis Sotware (Applied Biosystems).

*ZFTA-RELA* fusion transcript was investigated also by TaqMan real-time PCR using primers 5’-GCCGGTGTCCCAGCTT-3’ (forward), 5’-GCTCTGCCGGGAAGATGAG-3’ (reverse), 5’-CAAGGGCCCAGAACTG-3’ (probe sequence) in Applied Biosystems 7500 Fast DX Real-Time PCR Instrument. Housekeeping HPRT TaqMan gene assay was used as endogenous control.

In the BIOMECA reference laboratory in Bonn, the presence of *ZFTA-* and *YAP1-MAMLD1* fusions was examined by RT-PCR after RNA extraction from FFPE tumour probes using the AllPrep DNA/RNA FFPE kit (Qiagen, Venlo, The Netherlands) as previously described^24,25^. Further molecular analysis of gene fusions was implemented by using the Nanostring fusion panel which interrogates 88 recurrent fusions present in brain tumours, including the four most frequent *ZFTA*- (*C11orf95-RELA*) and two *YAP1*- (*YAP1-MAMLD1* and *YAP1-FAM118B*) fusions. ZFTA-like classified cases were examined further with Next-generation mRNA gene fusion panel using the TruSight Fusion Panel (Illumina, San Diego, CA, USA) as previous described^7^. Sequencing data were analysed by the Arriba tool (https://github.com/suhrig/arriba)^8^.
