## Supplemental Table 1 for "Optimising biomarkers for accurate ependymoma diagnosis, prognostication and stratification within International Clinical Trials: A BIOMECA study"

| Marker | Methodology | Centre/s | | |
|---|---|---|---|---|
| | | 1 | 2 | 3 |
| 1q gain | FISH | CBTRC, UK | CHSA, France | |
| | MLPA/MIP | CBTRC, UK | BONN, Germany | |
| | EPIC 850K methylation array | CBTRC, UK | DKFZ, Germany | |
| *ZFTA*- fusion | RT-PCR/Sequencing/Nanostring | BONN, Germany | COMO, Italy | |
| | IHC | BONN, Germany | COMO, Italy | |
| | FISH | CHSA, France | | |
| *YAP1*- fusion | RT-PCR/sequencing | BONN, Germany | COMO, Italy | |
| | IHC | BONN, Germany | COMO, Italy | |
| | FISH | CHSA, France | | |
| CDKN2A | MLPA/MIP | CBTRC, UK | BONN, Germany | |
| H3K27me3 | IHC | CBTRC, UK | BONN, Germany | VIENNA, Austria |
| Tenascin C (TNC) | IHC | CHSA, France | COMO, Italy | |

**Supp. Tbl 1: BIOMECA centre and methodology**
