## Supplemental Table 3 for "Optimising biomarkers for accurate ependymoma diagnosis, prognostication and stratification within International Clinical Trials: A BIOMECA study"

| | **ST-YAP1** | **ST-ZFTA** | **PFA** | **PFB** | **Other** |
|---|---|---|---|---|---|
| **Group** | ST-YAP1 | ST-ZFTA | PFA | PFB/PF-SE | SP-MPE<br>ST-PLAGL1<br>Neuroepithelial tumor with PATZ1-fusion |
| **Subgroup** | | ST-ZFTA<br>ST-ZFTA like | PFA-1<br>PFA-2 | PFB 1 - 3<br>PFB-4 | |
| **Subtype** | | ST-ZFTA<br>ST-ZFTA, Cl. 1<br>ST-ZFTA, Cl. 2<br>ST-ZFTA, Cl. 3<br>ST-ZFTA, Cl. 4 | PFA-1a<br>PFA-1b<br>PFA-1c<br>PFA-1d<br>PFA-1e<br>PFA-1f<br>PFA-2a<br>PFA-2b<br>PFA-2c | PFB-1<br>PFB-2<br>PFB-3<br>PFB-4 | |

**Suppl. Tbl. 3:** A layered diagnostic approach to molecular ependymoma classification
