## Supplemental Table 4 for "Optimising biomarkers for accurate ependymoma diagnosis, prognostication and stratification within International Clinical Trials: A BIOMECA study"

| Anatomical Location | Molecular Classification (DNA Methylation Array) | n = |
|---|---|---|
| Posterior Fossa (n= 111) | Posterior fossa ependymoma group A | 95 |
| | Posterior fossa ependymoma group B & posterior fossa subependymoma | 8 |
| | Score <0.9 | 8 |
| Supratentorial (n= 32) | Supratentorial ependymoma, ZFTA-fused | 18 |
| | Supratentorial ependymoma, ZFTA-like | 4 |
| | Supratentorial ependymoma, YAP1-fused | 1 |
| | Supratentorial ependymoma [non-defined type]; ST-PLAGL1 | 2 |
| | Posterior fossa ependymoma group A | 1 |
| | Posterior fossa ependymoma group B & posterior fossa subependymoma | 2 |
| | Score <0.9 | 3 |
| | Not classifiable | 1 |
| Spinal (n= 4) | Myxopapillary ependymoma | 2 |
| | Score <0.9 | 1 |
| | Not classifiable | 1 |
| *Total n =* | | *147* |

**Supp. Tbl. 4: Anatomical location and corresponding molecular classification**
