## Supplemental Figure 1 for "Optimising biomarkers for accurate ependymoma diagnosis, prognostication and stratification within International Clinical Trials: A BIOMECA study"

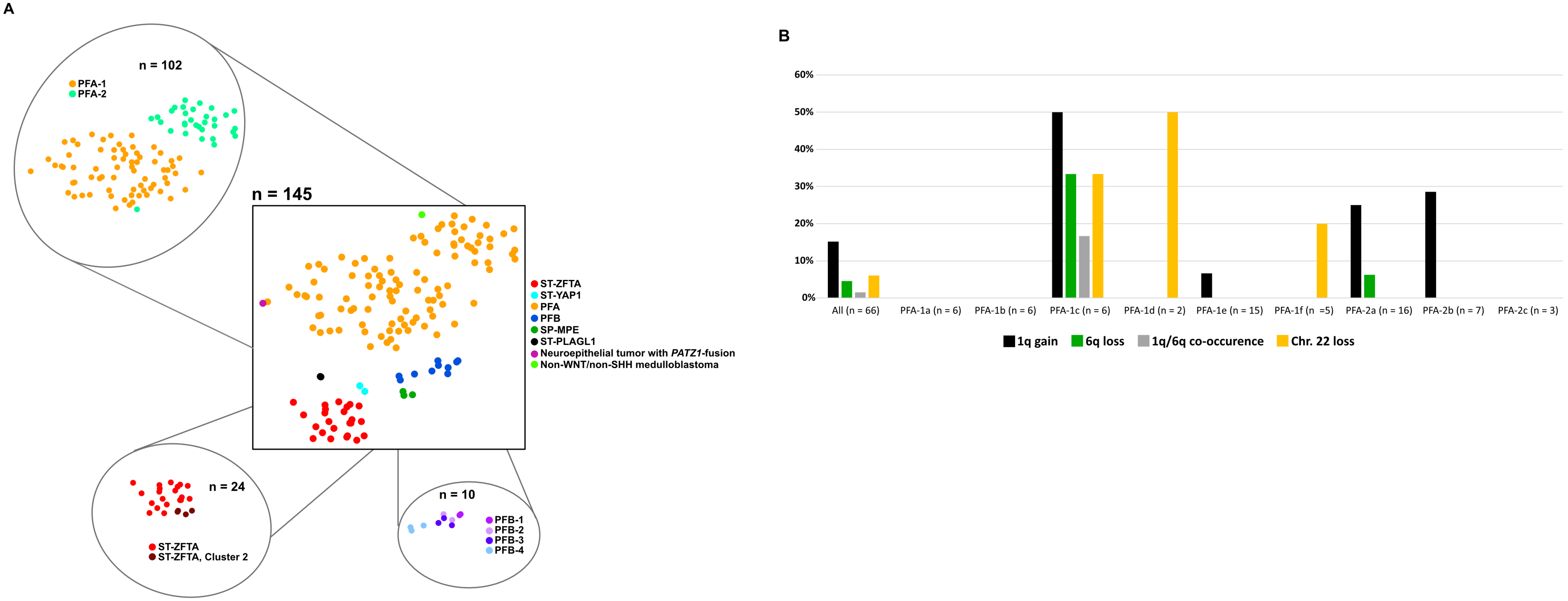


**Suppl. Fig. 1: Methylation based classification and copy number variation analysis for PFA subtypes**
A) t-SNE plot visualizing DNA-methylation based clustering of the BIOMECA cohort. Central t-SNE: Molecular groups, satellite t-SNEs: molecular subgroup for PFA, molecular subtype for ST-ZFTA and PFB. Samples are colorized based on the best available prediction regardless of the respective score. B) Bar chart depicting the relative frequency of group specific CNVs within PFA subtypes. The number of samples per subtype is given in parentheses.
