## Supplementary figures and images for "Optimising biomarkers for accurate ependymoma diagnosis, prognostication and stratification within International Clinical Trials: A BIOMECA study"

### Supplemental Figure 2

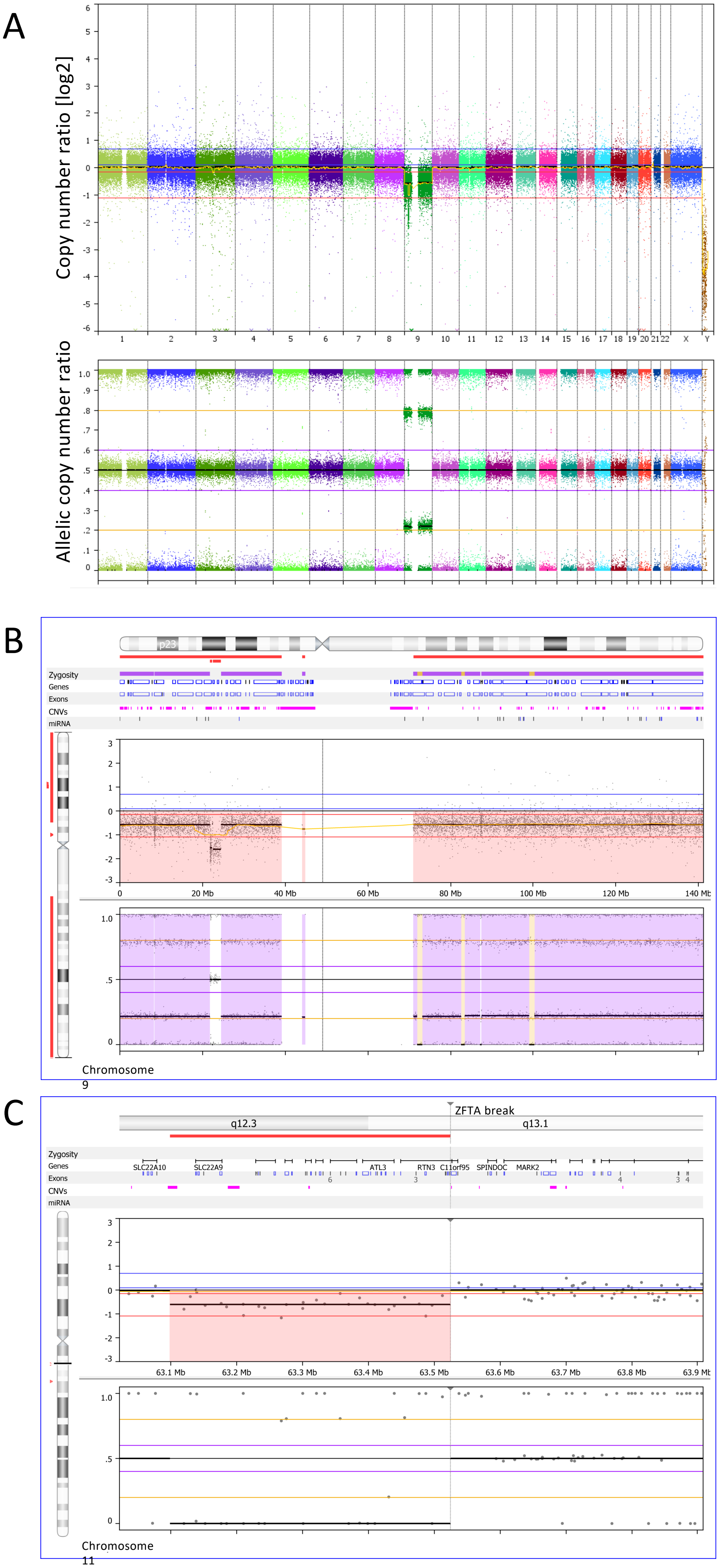


**Suppl. Fig. 2: Homozygous CDKN2A deletion**

### Supplemental Figure 3

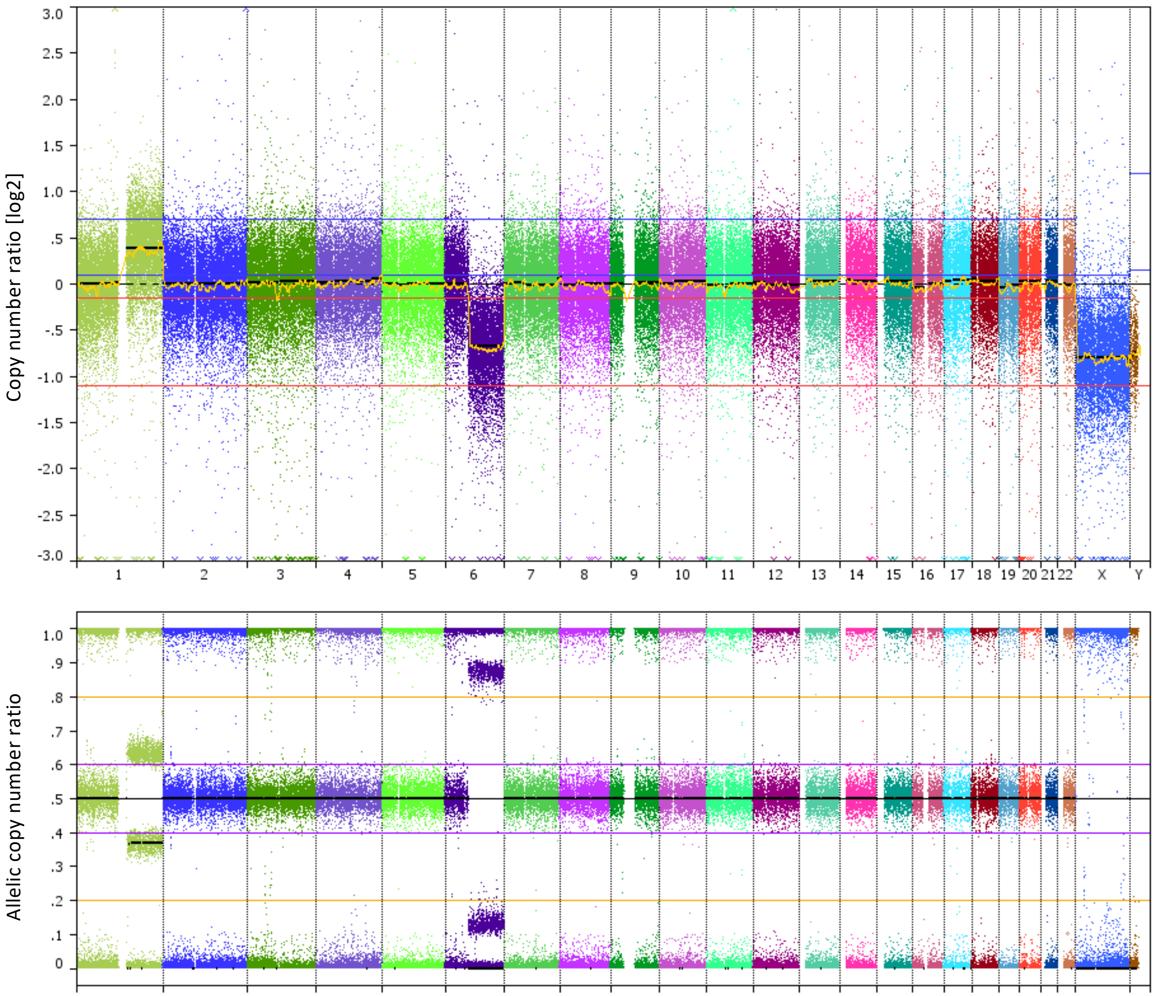


**Suppl. Fig. 3: 1q gain/6q loss co-occurrence by MIP**
