## Supplemental Figure 4 for "Optimising biomarkers for accurate ependymoma diagnosis, prognostication and stratification within International Clinical Trials: A BIOMECA study"

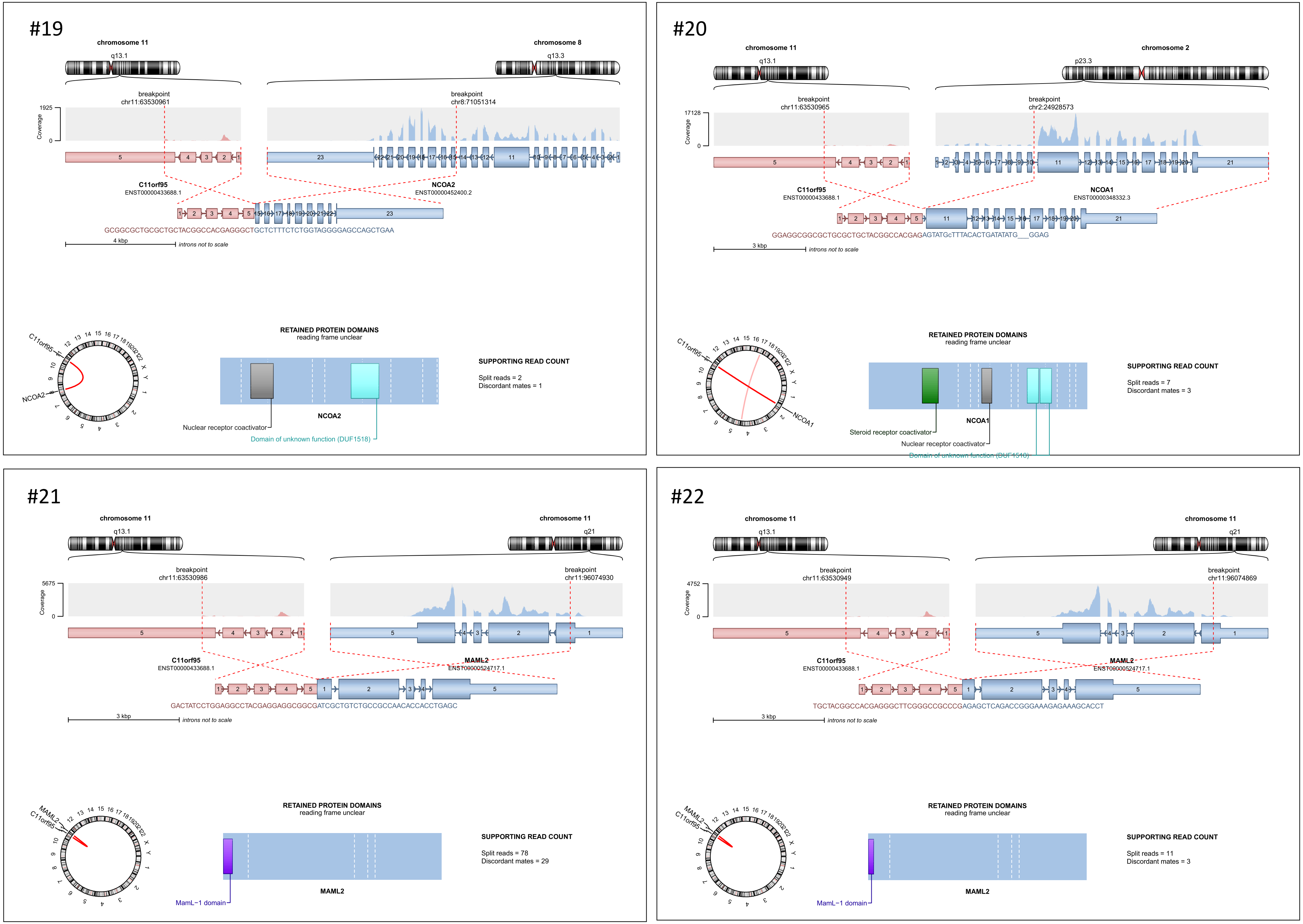


**Suppl. Fig. 4: RNA-sequencing of four ZFTA-like cases identified fusions of ZFTA-NCOA2 in #19, ZFTA-NCOA1 in #20, and ZFTA-MAML2 in #21 and #22.**
